# Inflammation, anemia and Vitamin D status determine infant thrombocytosis risk

**DOI:** 10.1101/2025.09.10.25335338

**Authors:** Sahaana S Rajagopalan, Brian M Dulmovits, Matthew Devine, Christopher S Thom

**Affiliations:** Division of Neonatology, Children’s Hospital of Philadelphia, Philadelphia, PA; Department of Pediatrics, University of Pennsylvania Perelman School of Medicine, Philadelphia, PA

**Keywords:** Platelet, NICU, thrombocytosis

## Abstract

Extreme thrombocytosis (EXT, >1000×10^3^ platelets/μl blood) occurs in infants due to infection, inflammation, and/or anemia. EXT can present diagnostic dilemmas, sometimes prompting invasive testing and anticoagulation therapy. Our prior worked identified heightened EXT rates in hospitalized infants compared with older children, but this analysis largely excluded ex-preterm infants at increased risk for infections and anemia – factors that promote thrombocytosis and EXT. The objectives of this study were 1) to define EXT rates, etiologies, and sequelae among infants hospitalized in tertiary neonatal intensive care units (NICUs) to assist clinical decision-making and 2) to ascertain factors that drive thrombocytosis and EXT in preterm and full-term patients. Retrospective analysis of thrombocytosis (>500×10^3^ platelets/μl) and EXT cases among 20,818 infants hospitalized in tertiary NICUs from 2011-2023 revealed thrombocytosis in 3% of all patients (8% of preterm infants). Both estimates were significantly lower than the incidence of thrombocytosis in pediatric patients in a quaternary NICU (15%). EXT rates were also markedly diminished in our tertiary unit (0.08% vs 0.5% in our quaternary NICU). Thrombocytosis was associated with leukocytosis and relative anemia, but not with thrombotic or bleeding complications. Vitamin D deficiency can drive thrombocytosis in adults and Vitamin D level was inversely corelated with platelet count among infants with thrombocytosis. Our findings suggest that Vitamin D supplementation among ex-preterm infants reduces thrombocytosis and EXT incidence, as opposed to developmental differences and/or organ immaturity in these patients. These results provide important context for clinical interpretations and responses to thrombocytosis in the preterm infant population.

## Introduction

Thrombocytosis (>500×10^3^ platelets/μl blood) and extreme thrombocytosis (EXT, >1000×10^3^ platelets/μl blood) result from infection, inflammation, anemia, hemolysis, and/or impaired platelet clearance in pediatric patients [1–4]. Primary thrombocytosis can also occur in the context of primary myeloproliferative disorders, but this is virtually nonexistent in pediatric patients [1]. High platelet counts can raise clinical concern for thrombosis, prompting anticoagulation therapy and invasive diagnostic testing [1]. Thus, understanding etiologies and sequelae of thrombocytosis and EXT are important to define differential diagnoses and potentially limit unnecessary tests and treatments. Understanding thrombocytosis etiologies can also help infer developmental differences in systems supporting production of megakaryocytes and platelets (megakaryothrombopoiesis), which change in early life [5,6].

Among pediatric patients at a quaternary hospital, EXT cases were frequently multifactorial, occurring secondary to inflammation, infection, iron deficiency, and/or splenic dysfunction [1,2]. EXT occurred more frequently in the context of critical illness, reflecting more severe pathophysiology. Interestingly, this condition was more common among infants <1 year of age, suggesting that altered megakaryothrombopoiesis in infants vs older children could underlie these observations. Indeed, neonatal megakaryocytes are less polyploid in neonatal bone marrow compared with older children and adults [6]. Neonatal megakaryocytes also have increased proliferation, but each megakaryocyte produces fewer platelets than adult bone marrow megakaryocytes [6]. Additionally, infant platelets differ from older individuals [7,8], potentially reflecting developmental changes in the types of megakaryocytes that produce platelet [9,10].

Infants born prematurely are at risk for infection, nutritional deficiencies, anemia, and organ immaturity that could underlie differences in propensities to experience thrombocytosis and/or EXT [11]. For example, premature infants are at increased risks of infection and often experience prolonged periods of critical illness requiring respiratory and cardiovascular support. In general, this clinical context should place premature infants at heightened risk for thrombocytosis and EXT. However, prior studies of neonatal and pediatric thrombocytosis have largely neglected inborn infants hospitalized in tertiary neonatal intensive care units (NICUs), which care for most premature infants born at <37 weeks gestational age.

We therefore sought to understand how our prior findings in a quaternary NICU population would translate to infants hospitalized in level 3 NICUs in our hospital network. The aims of this study were to clarify the prevalence, etiologies, and sequelae of thrombocytosis, including thrombotic complications of EXT in preterm infants that may warrant antiplatelet therapy.

Retrospective evaluation of 20,818 infants over more than 10 years revealed that thrombocytosis and EXT were rare and dramatically decreased vs infants hospitalized at our quaternary hospital. These findings were consistent in a focused analysis of premature infants that revealed thrombocytosis and EXT as uncommon but warranting for evaluation for infection or inflammation.

## Methods

### Thrombocytosis and EXT case identification

We retrospectively identified and profiled 20,818 cases of thrombocytosis (>500×10^6^ platelets/μl) among infants hospitalized in two tertiary NICUs in our hospital network from 2011-2023. Infants were predominantly delivered at these respective NICUs, permitting a global assessment of the neonatal hospitalization course. Statistical comparisons and trends were similar when analyzing all blood counts or restricting analyses to unique patients. We opted to include all blood counts given the possibility that some patients had recurrent thrombocytosis, which happened infrequently. We manually reviewed all cases of EXT (>1000×10^3^ platelets/μl) and near-EXT (>900×10^3^/μl) to determine etiologies and clinical sequelae. This study was approved by the University of Pennsylvania and Children’s Hospital of Philadelphia Institutional Review Boards.

### Statistical analysis and plotting

We used R (v4.2.3) and GraphPad Prism (v10) for statistical analysis and graphical plotting.

### Data availability

Summary data are available upon request.

## Results

### Thrombocytosis is rare among preterm infants in a tertiary NICU

In a retrospective analysis of 20,818 hospitalized infants at two neonatal intensive care units (NICUs), we identified a total of 713 infants (3%) with thrombocytosis (>500×10^3^/μl) and 3 cases of EXT (>1000×10^3^/μl, **Figure 1A**). No infants were anticoagulated in response to EXT, and there were no thrombotic or bleeding complications directly linked to instances of EXT.

**Figure 1.**
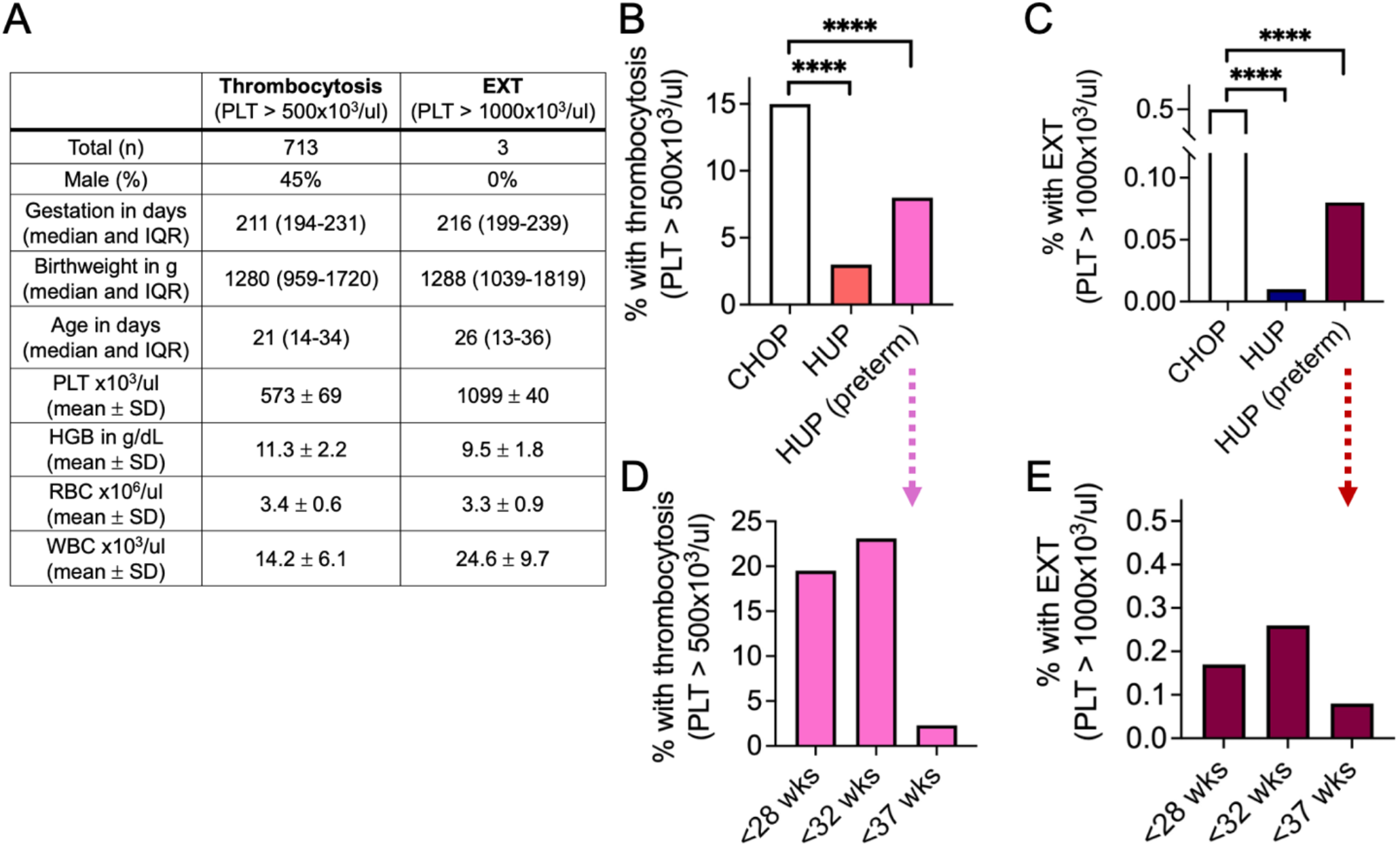
Thrombocytosis and extreme thrombocytosis (EXT) rates among infants hospitalized at a tertiary vs quaternary care units, including breakdown by gestational age for preterm infants. **A**) Blood counts and demographics for infants with thrombocytosis or EXT in our cohort of 20,818 total infants. Blood counts represent values at the time of thrombocytosis or EXT. PLT, platelet count. HGB, hemoglobin. RBC, red blood cell count. WBC, white blood cell count. **B**) Thrombocytosis rates among infants hospitalized at our tertiary care unit, including a breakdown of those both preterm (<37 weeks gestation at birth). Reference comparison to historical data from a quaternary referral NICU [1]. **C**) EXT rates among infants hospitalized at our tertiary care unit, including a breakdown of those both preterm (<37 wks gestation at birth). Reference comparison to historical data from a quaternary referral NICU [1]. **D**) Breakdown of thrombocytosis rates based on degree of prematurity. **E**) Breakdown of EXT rates based on degree of prematurity.

These rates of thrombocytosis and EXT observed in this population are significantly less compared to pediatric patients hospitalized in a quaternary hospital [1] (**Figure 1B-C**). This was surprising, given high rates of sepsis, inflammation, and anemia among this cohort, which we have previously shown to drive pediatric thrombocytosis [1].

Many premature infants experience critical illness during prolonged hospitalizations, leading us to hypothesize that increased EXT rates might be higher among extremely preterm infants and/or those with lowest birth weights. Indeed, thrombocytosis and EXT rates were relatively increased among smaller and more premature infants (**Figure 1D-E**). However, even among the most premature subset, EXT rates did not reach the levels previously observed among pediatric patients in a quaternary hospital (χ^2^<0.001).

### Thrombocytosis among preterm infants reflects inflammation

Multifactorial etiologies often underlie thrombocytosis and EXT, most notably including infection, inflammation, and anemia [1], as well as organ dysfunction related to platelet clearance [2]. To disentangle factors related to thrombocytosis, we evaluated clinical associations between platelet counts, blood traits, and other analytes (**Figure 2A**). We noted significant positive associations between platelet counts and white blood cell counts, indicating that thrombocytosis occurred with leukocytosis, most likely in the setting of infection and/or inflammation (**Figure 2B**).

**Figure 2.**
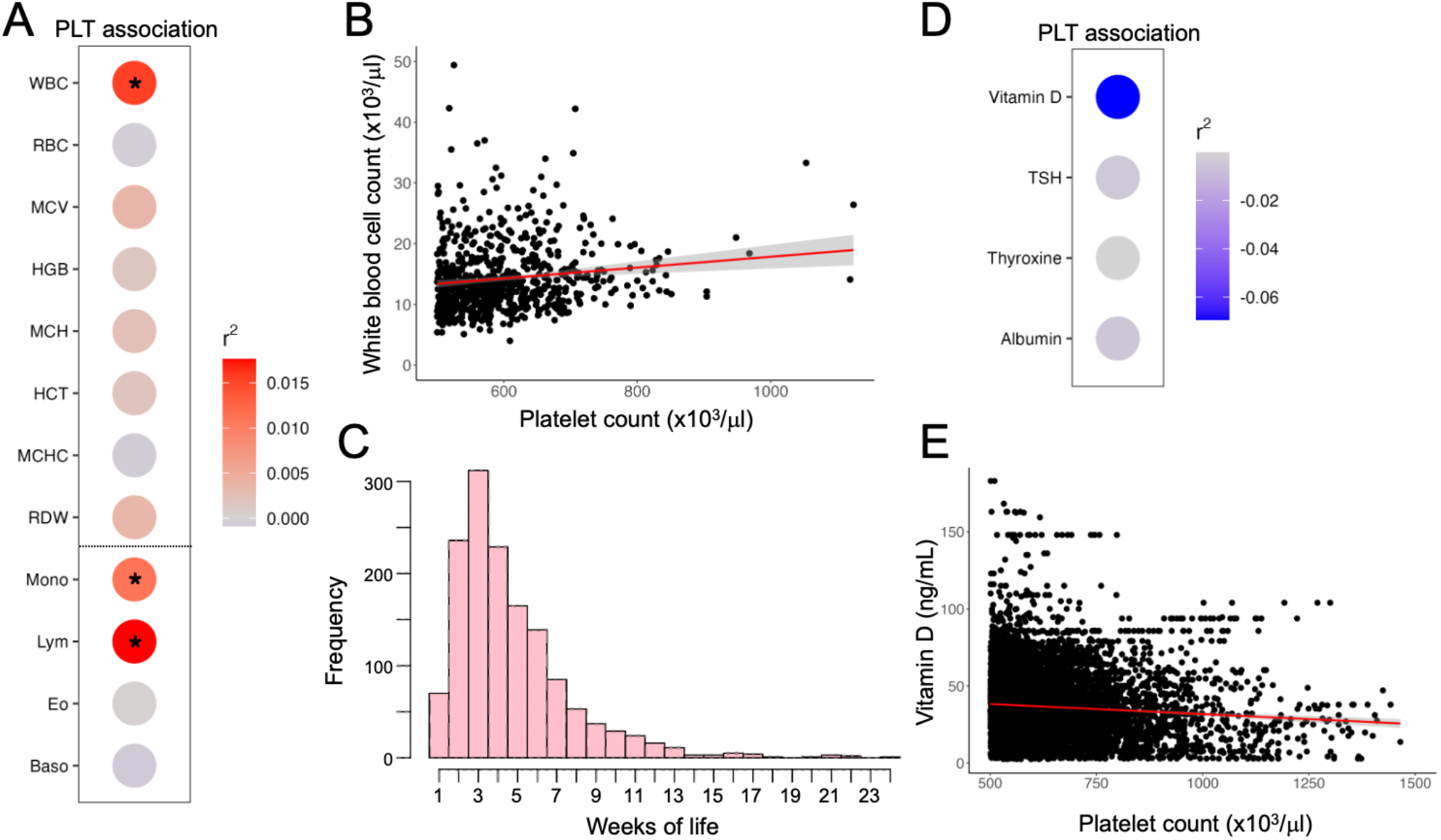
Platelet count is associated with altered white blood cell count and Vitamin D level, with thrombocytosis occurring most frequently around 3-4 weeks of life in temporal relation with hemoglobin nadir. **A**) Heat map depicting r^2^ values with platelet count (PLT) associations for the indicated traits. Traits with significant correlation, as defined by a line of best fit slope that deviates significantly from 0 are indicated with an asterisk [Pr(>|t|)<0.05]. Red dots indicate positive correlation and blue dots indicate negative (inverse) correlation. A breakdown of quantitative leukocyte parameters is shown below the dashed line, with Monocyte (Mono) and Lymphocyte (Lym) counts showing significant direct association with PLT count. **B**) Scatterplot comparing platelet count vs white blood cell count in unique infants with thrombocytosis (maximum 1 dot per infant). Line of best fit is shown in red with standard deviation in gray. Pr(>|t|)=1.8x10^-2^, meaning the slope of this line significantly differs from 0. **C**) The incidence of thrombocytosis peaks in the 3^rd^ week of life in this patient cohort. This histogram includes episodes of recurrent thrombocytosis that occurred in some patients. **D**) Heat map depicting r^2^ values with platelet count (PLT) associations for the indicated traits. Blue dots indicate negative (inverse) correlation. **E**) Scatterplot comparing platelet count vs Vitamin D level in infants with thrombocytosis, including patients in tertiary and quaternary NICUs. Line of best fit is shown in red with standard deviation in gray. Pr(>|t|)=2.9x10^-11^, meaning the slope of this line significantly differs from 0.

### Thrombocytosis incidence peaks during the physiologic hemoglobin nadir

Interestingly, there was not a significant association between platelet count and red cell parameters (e.g., hemoglobin) among thrombocytosis cases (**Figure 2A**). This may suggest that anemia was not a major contributing factor to the degree of thrombocytosis among this cohort, although prior transfusions may confound this interpretation. However, the hemoglobin levels observed among this cohort revealed relative anemia compared with typical expectations (**Table 1**).

**Table 1.** Blood counts and clinical context for EXT cases among infants in tertiary neonatal intensive care units. DOL refers to the first instance of EXT or near-EXT; in some cases EXT was redemonstrated the subsequent day as well. One case (*) occurred in an infant with a congenital hemolytic anemia, which can predispose to thrombocytosis [3]. PLT, platelet count (x10^3^/μl). WBC, white blood cell count (x10^3^/μl). HGB, hemoglobin (g/dL). Normal ranges are shown.

| Case<br>(Normal range) | Birth gest. age<br>(range in weeks) | Day of life<br>(range in weeks) | <u>Counts at time of EXT</u> |  |  |
| --- | --- | --- | --- | --- | --- |
| | | | PLT<br>(150-400<br>$\times 10^3/\mu\text{l}$) | WBC<br>(8.0-15.4<br>$\times 10^3/\mu\text{l}$) | HGB<br>(12.5-20.5<br>g/dL) |
| 1* | 37-42 | 0-5 | 1124 | 26.4 | 9.3 |
| 2 | 32-37 | 0-5 | 948 | 21 | 10.9 |
| 3 | 27-32 | 5-10 | 1110 | 17.8 | 11.2 |
| 4 | 27-32 | 0-5 | 904 | 12.1 | 8.7 |
| 5 | 27-32 | 0-5 | 968 | 18.4 | 11 |
| 6 | 22-26 | 0-5 | 1053 | 33.3 | 7.9 |

We next assessed timing of thrombocytosis and EXT among this cohort to gain further insight into related physiology and etiologies. Thrombocytosis peaked around ∼1 month of life, at a time that often correlates with a hemoglobin nadir from physiologic anemia and anemia of prematurity [12] (**Figure 2C**). This finding supports relative anemia and/or subclinical iron deficiency as contributing factors to thrombocytosis among preterm infants, as in other cohorts [1].

### EXT occurs in multifactorial contexts among preterm infants

To elucidate specific clinical etiologies driving thrombocytosis, we profiled EXT cases among our cohort. Given the paucity of EXT cases, we extended the typical definition of EXT to include all cases of thrombocytosis with platelet counts exceeding 900×10^3^/μl (**Table 1**). These extreme cases highlighted concurrent leukocytosis and anemia in the setting of EXT, with our clinical case review indicating acute infection or inflammation that typically preceded the development of EXT. Interestingly, one case also occurred around birth in an infant with a congenital hemolytic disorder, which we and others have previously found to drive thrombocytosis in some cases [3]. All cases recovered within a week without direct clinical intervention. None incurred thrombotic complications.

### Vitamin D levels are inversely correlated with platelet count among infants with thrombocytosis

We then investigated etiologies for the low thrombocytosis rates observed among our cohort of infants vs full term infants from a quaternary hospital (**Figure 1**). We hypothesized that organ immaturity and/or developmental differences in organ function may be responsible. The liver primarily produces thrombopoietin (TPO) and acute phase reactants. Since these factors are not routinely measured in infants, we analyzed albumin levels as a proxy for liver function. Albumin levels were nearly identical among infants with thrombocytosis in our study cohort vs patients in our quaternary NICU, suggesting intact liver function (**Table 2**).

**Table 2.**
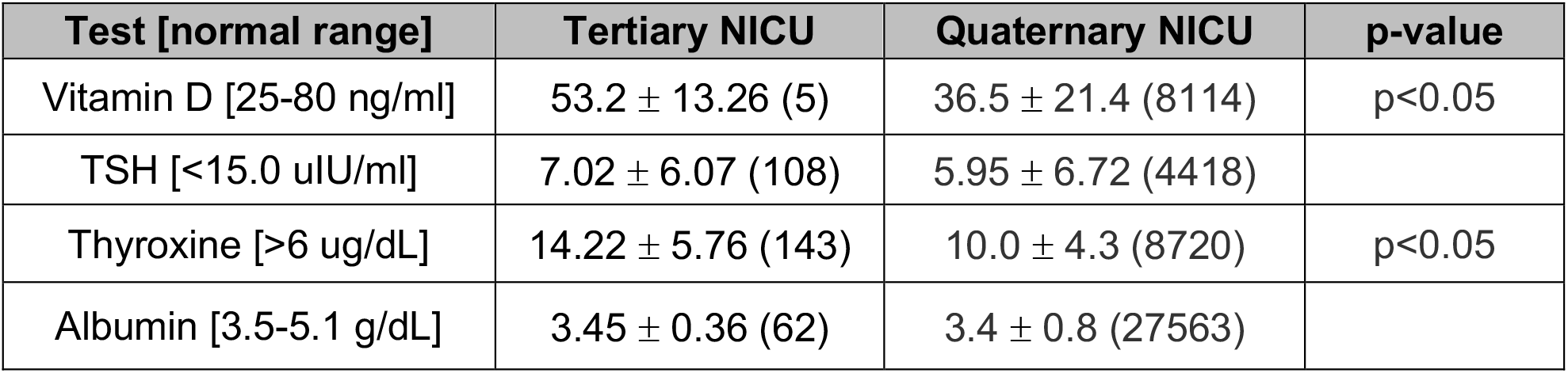
Select clinical test results in temporal relation to thrombocytosis in infants hospitalized at tertiary or quaternary NICUs. Statistical comparisons between populations were made by two-sided Welch’s t test. TSH, thyroid stimulating hormone. Statistically significant differences are indicated by p-values and labeled in bold. All parameters were associated with platelet count by linear regression in quaternary NICU patients (p<0.05), but were not significantly correlated with platelet counts in tertiary NICU patients. All values reflect mean ± standard deviation (n unique patients).

We then analyzed thyroid hormone levels among the cohorts, since hypothyroxinemia is common among preterm infants [13] and thyroid hormone levels are correlated with adult platelet counts [14]. Levels of thyroid stimulating hormone (TSH) and thyroxine were within normal range in our cohort and, interestingly, were elevated in this cohort when compared to infants from our quaternary hospital (**Table 2**). Thus, abnormal thyroid hormone levels do not appear responsible for the paucity of thrombocytosis and EXT among infants in our cohort.

We next analyzed Vitamin D levels in our cohort and quaternary NICU patients with thrombocytosis, given links between Vitamin D deficiency and thrombocytosis in adult populations [15,16]. Vitamin D level and platelet count are inversely correlated in adults, but this trend has not been established in pediatric patients. We identified an inverse correlation between Vitamin D level and platelet count in infants from our tertiary NICU, and a significant inverse correlation when we included infants from our quaternary NICU (**Figure 2D-E** and **Table 2**). We also noted that total Vitamin D levels were significantly increased in our tertiary NICU cohort (53.2±13.3 ng/mL) compared to quaternary NICU patients (vs 36.5±21.4 ng/mL, p<0.05, **Table 2**). Higher Vitamin D level among tertiary NICU patients would be expected to blunt inflammatory responses and thrombocytosis [17,18], despite clinical circumstances that should otherwise drive higher levels of thrombocytosis [1].

## Discussion

Among infants hospitalized at our tertiary care NICU, thrombocytosis resulted from a multifactorial combination of infection, inflammation, and relative anemia. This supports the notion that the predominance of pediatric thrombocytosis is a secondary response to an inciting etiology [1]. While thrombocytosis and EXT can cause significant clinical concerns, EXT did *not* cause thrombotic complication and resolved without intervention in all cases among the cohort examined in this study. Our findings show that any degree of thrombocytosis (e.g., >500-600×10^3^ platelets/μl) should warrant consideration and that higher levels of thrombocytosis (>1000×10^3^ platelets/μl) may not be physiologically supported in some premature infants.

While the etiologies and sequelae of thrombocytosis and EXT matched other pediatric patients [1], there was a relative paucity of EXT cases among our tertiary NICU patients (**Figure 1**).

Lower rates of EXT were particularly notable among preterm infants born extremely prematurely or with extremely low birth weight, who are at heightened risk for infection, anemia, and critical illness - factors linked to the development of EXT [1,11]. These findings are not overtly linked to liver or thyroid immaturity or developmental differences, although we could not exclude hepatic secretion of key platelet-related cytokines given a lack of routine clinical testing (e.g., IL6, TPO).

Our findings link Vitamin D deficiency with thrombocytosis in this cohort. Post-hoc analysis of a quaternary NICU patient cohort show an inverse correlation between Vitamin D level and platelet count (**Fig. 2E**). Comparatively, Vitamin D levels were higher in tertiary NICU patients. The nutritional status of premature infants in these tertiary NICUs is closely monitored and controlled, including robust Vitamin D supplementation. This may have blunted thrombocytosis responses and precluded development of EXT among these patients. We expect related mechanisms to include Vitamin D-dependent effects to limit inflammation and platelet activation, as have been shown in adults [17,18].

Megakaryocyte and platelet production differ in neonates compared with older children and adults, and these differences likely underlie increased rates of thrombocytosis and EXT among infants in some pediatric populations [1]. However, findings in the present study highlight the importance of exogenous systemic factors (e.g., Vitamin D levels) in determining platelet production, opening exciting avenues for future study. The effects of Vitamin D and/or other downstream systemic factors (e.g., inflammatory responses) may be harnessed to augment platelet production in vitro or in vivo to support novel cell therapeutic development and used to help understand processes that support megakaryothrombopoiesis in neonates and infants.

We do not anticipate that Vitamin D supplementation and relatively reduced propensity for thrombocytosis would have negative health effects. Immunologic roles for platelets are well established, including direct binding of bacteria for translocation to the spleen, immune cell recruitment, and the release of inflammatory cytokines from activated platelets [8,19–21]. However, there is no evidence to suggest that extremely high platelet levels (e.g., EXT with >1000×10^3^ platelets/μl) are superior from an immunologic perspective than more modest thrombocytosis (500-1000×10^3^ platelets/μl). Although platelets differ between newborns and adults [8], substantial changes occur during the first weeks of life that ‘normalize’ the hemostatic and immunologic environments [6,22,23]. Most thrombocytosis occurred after this transition in our cohort (∼3 weeks of life, **Fig. 2C**). These findings reinforce the importance of optimal nutrition to support normal organ function in preterm infants, in addition to rigorous assessment in the setting of thrombocytosis to determine clinical management.

## Conflicts of Interest

The authors declare no conflicts of interest in relation to this work.

## Acknowledgements

This study was supported by the National Institutes of Health (T32 HL007150 to BMD, K99 HL156052 to CST, R00 HL177827 to CST) and the Children’s Hospital of Philadelphia.

## Notes

### Competing Interest Statement

The authors have declared no competing interest.

### Author Declarations

The Institutional Review Board of the University of Pennsylvania gave ethical approval for this work. The Institutional Review Board of the Childrens Hospital of Philadelphia gave ethical approval for this work.

